# Exploring the Impact of Oral Health and Vaccination on Pneumonia-causing Bacteria: Insights from Predictive Modeling

**DOI:** 10.1101/2025.05.23.25328168

**Authors:** Ryann N. Whealy, Skylar Timm, Tara N. Furstenau, Alexander Roberts, Sara Maltinsky, Sydney Wells, Kylie Drake, Kayla Ramirez, Candice Bolduc, Ann Ross, Talima Pearson, Viacheslav Y. Fofanov

**Affiliations:** Pathogen and Microbiome Institute, Northern Arizona University, Flagstaff, Arizona, United States of America; School of Informatics, Computing, and Cyber Systems, Northern Arizona University, Flagstaff, Arizona, United States of America; Mobile Dentistry of Arizona, Mesa, Arizona, United States of America

## Abstract

Nursing home acquired pneumonia (NHAP) is a leading cause of mortality in long-term care facilities (LTCFs). The primary mechanism of infection in older individuals is macro-aspiration of opportunistic pathogens colonizing the oral and nasal cavities. Pneumococcal vaccination is the primary preventative measure, but oral hygiene is increasingly explored as a strategy for reducing oral colonization. While promising, oral health interventions have shown mixed results, likely due to variation in the way that different NHAP-associated pathogens respond to these interventions. To test this, we analyzed oral health survey responses, pneumococcal vaccination status, and longitudinal colonization to identify factors linked to carriage of each pathogen. We found that oral hygiene impacted carriage of bacteria which more frequently colonized the oral cavity (*Haemophilus influenzae* and *Streptococcus pneumoniae*), but had limited effect on bacteria that were more prevalent in the anterior nares (*Staphylococcus aureus* and *Pseudomonas aeruginosa*). Pneumococcal vaccination reduced oral colonization by both *S. pneumoniae* and *H. influenzae*, suggesting community level interactions. Our results suggest that oral hygiene interventions are likely to only be effective against pathogens that primarily colonize the oral cavity, underscoring the importance of identifying the primary reservoirs of NHAP-associated pathogens when developing effective intervention strategies.

## Introduction

Pneumonia is a common lower respiratory tract infection, accounting for ~4.9 million cases annually and $13.4 billion in healthcare costs in the US [1]. Older adults (≥ 65 years old) account for about 24% of cases [1], 30% of whom require hospitalization [2], with mortality ranging from 13-41% post-admission [3]. Nursing Home-Acquired Pneumonia (NHAP) has a higher mortality rate than Community-Acquired Pneumonia (CAP) and is 8-11 times more likely to affect elderly patients [4–8]. NHAP and CAP are commonly caused by aspiration of upper respiratory tract colonizing bacteria such as *S. pneumoniae, S. aureus, H. influenzae*, and *P. aeruginosa [9–19]*, making carriage of these pathogens a major risk factor for infection [17,18,20]. Aspiration pneumonia accounts for up to 86% of pneumonia cases in older adults [21], and approximately 10% of aspiration events lead to pneumonia in those aged 80 and older [22].

Vaccination remains the primary strategy for pneumonia prevention. At the time of this study, the CDC recommended the pneumococcal vaccine which protects against *S. pneumoniae* for anyone over 65 and all long-term care facility (LTCF) residents [23]. Conjugate vaccines reduce vaccine-type pneumococcal pneumonia and invasive disease [24,25], and have lowered carriage of covered serotypes, though non-vaccine serotypes are emerging to fill this void [26–29]. Despite their effectiveness, pneumococcal vaccines are not designed to protect against other pneumonia-causing pathogens, and age-related immune decline leaves older adults at continued risk. Additional prevention strategies are needed, especially for those who are unable or unwilling to be vaccinated.

Poor oral hygiene, which is common in elderly populations [30], can increase the prevalence of these colonizing opportunistic pathogens (COPs) in dental plaque [31,32] and raise the risk of aspiration pneumonia [33,34]. While some trials report reductions in pneumonia-related symptoms and mortality with oral care [35–39], others suggest a weak or non-significant effect [40–42]. These inconsistencies may reflect differences in pathogen reservoirs as oral hygiene is likely more effective against pathogens primarily colonizing the oral cavity than those which primarily reside in the nares (e.g. *S. aureus* and *P. aeruginosa*) [43], or against infections driven by direct viral or bacterial transmission [40]. The diversity of pneumonia-causing COPs, combined with differences in their anatomical reservoirs presents a significant challenge in determining which pathogens are likely to respond to oral care interventions [44]. This study aims to address this gap by examining how past and current oral care impacts carriage of *H. influenzae, P. aeruginosa, S. pneumoniae*, and *S. aureus*.

## Main text

### Methods

#### Study participants and data collection

Residents from three LTCFs in the Phoenix, AZ metropolitan area were enrolled in a two-year study (Northern Arizona University Institutional Review Board #1766728). Demographic information (race, ethnicity, age, and sex) and pneumococcal vaccine status was collected from each participant. A multiple-choice oral health care questionnaire was distributed to all participants (Table 1).

**Table 1:**
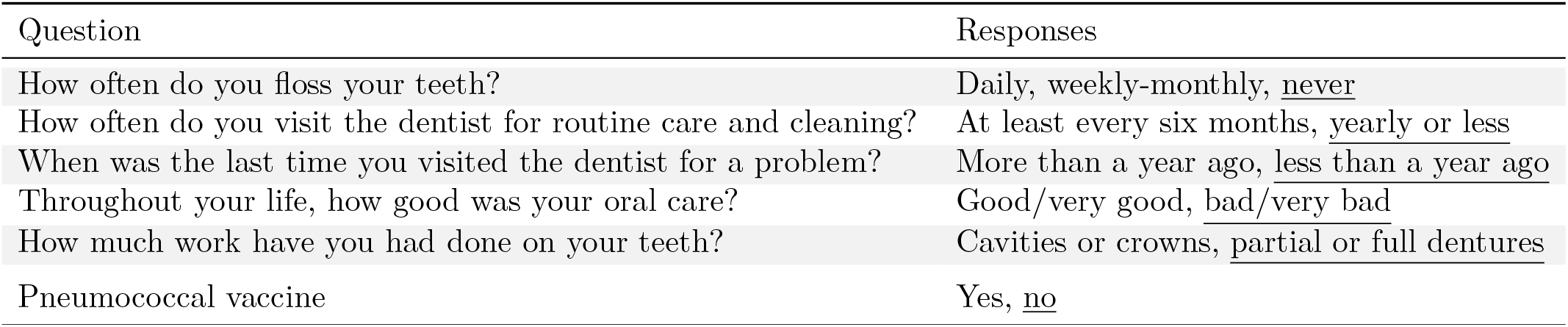
Participant survey questions and responses. The response category used as a reference in logistic regressions is underlined.

| Question | Responses |
| --- | --- |
| How often do you floss your teeth? | Daily, weekly-monthly, <u>never</u> |
| How often do you visit the dentist for routine care and cleaning? | At least every six months, <u>yearly or less</u> |
| When was the last time you visited the dentist for a problem? | More than a year ago, <u>less than a year ago</u> |
| Throughout your life, how good was your oral care? | Good/very good, <u>bad/very bad</u> |
| How much work have you had done on your teeth? | Cavities or crowns, <u>partial or full dentures</u> |
| Pneumococcal vaccine | Yes, <u>no</u> |

#### Sample collection and processing

Samples were collected biweekly through supervised self-swabbing of the anterior nares and oral cavity. Samples were placed in liquid Amies transport media and kept on ice prior to storage at −20°C. DNA was extracted from liquid Amies and tested for *H. influenzae, P. aeruginosa, S. pneumoniae*, and *S. aureus* using a multiplexed qPCR [43].

#### Data analysis

To address multicollinearity among oral health survey responses and improve statistical power, we applied principal component analysis (PCA) to reduce the dimensionality of the input variables (R v4.4.1); all variables were centered and scaled prior to analysis. Binomial logistic regression was used to predict the proportion of weeks where participants tested positive for each pathogen in oral samples. Each model included the top two principal components and pneumococcal vaccination status as main effects.

## Results

### Participant demographics and oral carriage

The study included 121 participants, with 50 completing the oral health survey. Demographic comparison confirmed similar characteristics between respondents and the overall cohort (Figure 1A). Respondents’ median age (82) closely matched the overall median (83), though respondents had a higher proportion of men (32% vs. 24%) and slightly lower pneumonia vaccination rates (70% vs. 73.6%). These distributions reflect demographics of Arizona LTCF residents, suggesting that our sample is representative of this population [45]. The proportion of sampling events that participants tested positive for each pathogen exhibited skewed distributions suggesting prolonged colonization in a subset of individuals (Figure 1B). Similar patterns appeared among survey respondents.

**Figure 1:**
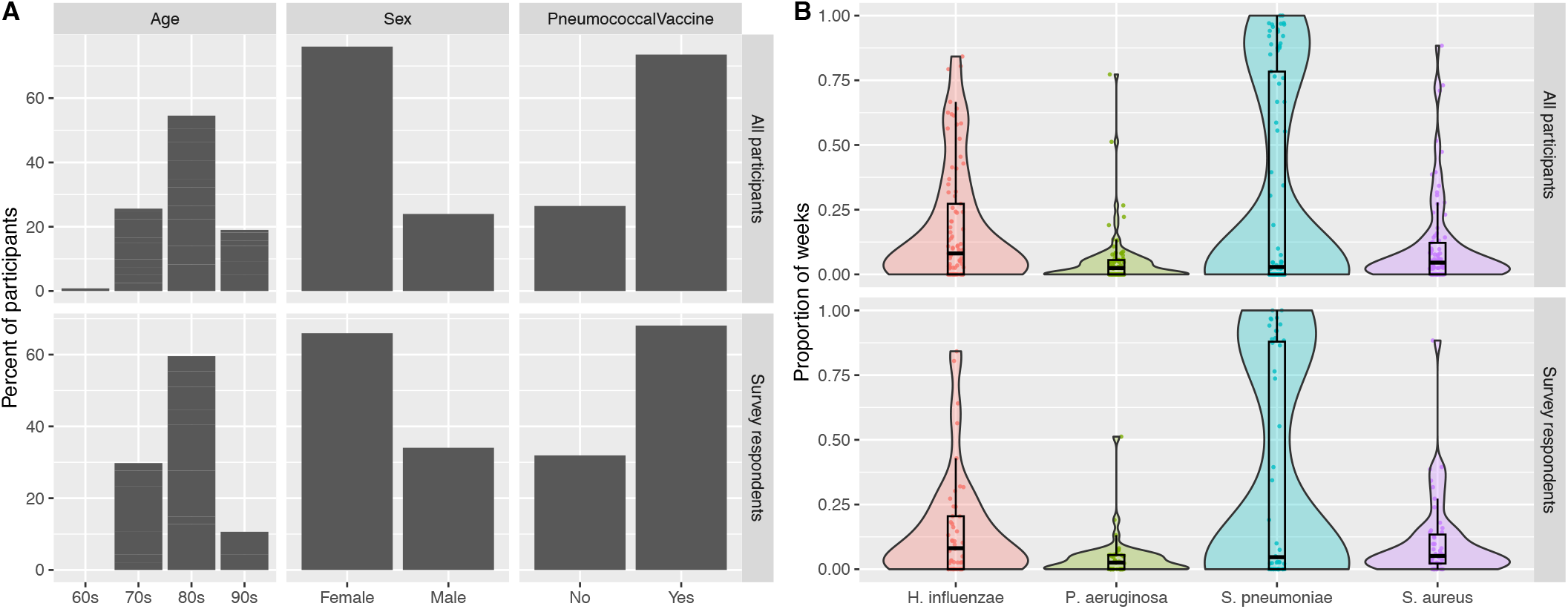
Participant Demographics and duration of oral carriage. A) Comparison of demographics between all study participants (n=121) and the subset of participants who responded to the oral health survey (n=50). B) Comparison of the proportion of sampling events where pathogens were detected from oral swabs in the total dataset versus the subset who responded to our oral health survey, among those who attended at least ⅓ of sampling events. Patterns of persistent colonization by *S. pneumoniae* were detected in oral samples.

### Summarizing oral health behaviors using PCA

To address high correlation among oral health behaviors, we used PCA to reduce dimensionality and improve interpretation. PC1 (36.7% of the total variance) captured chronic oral health behaviors — specifically, frequency of flossing, self-reported quality of lifetime oral care, and the extent of past dental work (loadings > 0.5). PC2 (26.5% of the variance) reflected recent or acute dental care behaviors, most notably routine dental visit frequency (loading = 0.65) and time since last treatment for a dental issue (loading = −0.69). Combined, PC1 and PC2 captured 63.2% of the total behavioral variance (Figure 2), and were used as oral health behavior summary predictors of colonization risk.

**Figure 2:**
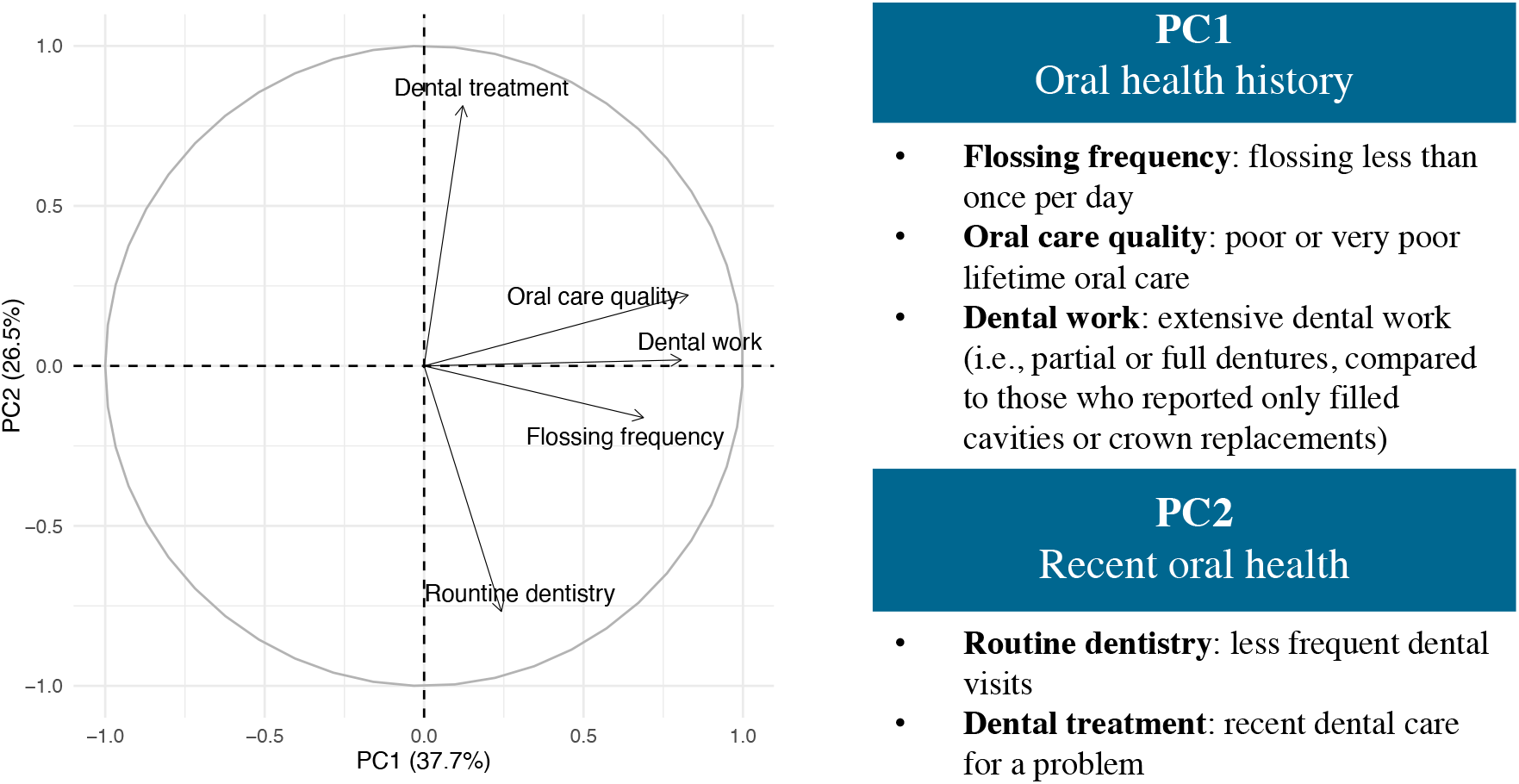
PCA biplot. Relationship of oral health survey variables to each other and principal components. Arrow length shows how much each variable contributes to the principal components, with longer arrows representing a stronger contribution, and arrow direction indicating the direction of correlations.

### Oral health behaviors and oral colonization persistence

Chronic oral health deficits (PC1), including not flossing regularly, poor lifetime oral hygiene, and extensive dental work, increased odds of more persistent colonization with *H. influenzae* (10.2%, P = 0.08) and *S. pneumoniae* (20.4%, P < 0.001). Acute or reactive oral care behaviors (PC2), characterized by less frequent dental visits or dental visits primarily driven by immediate problems, also predicted increased persistent colonization odds for these same pathogens (*H. influenzae* (17.3%, P < 0.01) and *S. pneumoniae* (9.7%, P = .05)). Neither acute nor reactive oral care behaviors significantly influenced colonization persistence by *P. aeruginosa* or *S. aureus* (P > 0.3).

### Effects of Pneumococcal vaccination

Pneumococcal vaccination was a significant protective factor against persistent oral colonization by *H. influenzae* and *S. pneumoniae*, reducing colonization odds by 56.5% (*P* < 0.01) and 48.29% (*P* < 0.01), respectively. In contrast, vaccination had no significant effect on *S. aureus* and was associated with a 143.82% increase in *P. aeruginosa* colonization odds (P<0.01). In the full study population (including participants who did not complete the oral health survey), these results were largely consistent. The odds remained significantly reduced for *H. influenzae* (18.86%, *p* = 0.05) and *S. pneumoniae* (27.17%, *p* < 0.001) and remained significantly increased for *P. aeruginosa* (84.15%, *p* < 0.01), however there was also a significant increase in odds for *S. aureus* (37.29%, *p* < 0.05).

## Discussion

This study aimed to identify modifiable factors that influence oral colonization of four NHAP-associated pathogens in elderly people. Understanding how oral care habits and vaccination relate to colonization can inform more effective preventive strategies and clinical practices.

As expected, pathogens more frequently colonizing the oral cavity (*H. influenzae* and *S. pneumoniae*) were impacted by oral health behaviors. *S. pneumoniae* carriage persistence was significantly reduced in participants that had better chronic oral hygiene behaviors (i.e., increased flossing, good lifetime oral care, and less extensive dental work). *H. influenzae* carriage was significantly reduced in participants who visited the dentist more frequently for cleanings and were less reactive in seeking care for an immediate problem. In contrast, *P. aeruginosa* and *S. aureus* colonization appeared unaffected by oral hygiene behaviors. This aligns with our previous findings that these bacteria primarily colonize the nasal cavity rather than the oral cavity [43].

Pneumococcal vaccination significantly reduced oral colonization by *S. pneumoniae* and *H. influenzae*, and was associated with lower overall and more transient *S. pneumoniae* carriage in both the subset of participants that responded to the oral health survey and in the full study population. These results are consistent with evidence that the pneumococcal conjugate vaccine prevents disease by reducing the likelihood of *S. pneumoniae* acquisition and carriage [46]. Previous reports have found an increase in *H. influenzae* carriage after pneumococcal vaccine administration [47], however others have found an inverse relationship [48], consistent with the findings of this study. Conversely, vaccinated individuals in our study had higher colonization rates of *P. aeruginosa* and *S. aureus*, consistent with previous reports that suggest these species may either compete with *S. pneumoniae* [49–51] or that broader shifts in the microbial composition following vaccination may impact their carriage[47,52]. These results highlight the importance of considering how interventions targeting one pathogen may impact others in the microbial community [52].

Although improved oral hygiene appears beneficial, effects vary by pathogen, suggesting that the success of interventions may depend on the organisms involved. Most prior studies did not differentiate between etiological agents when evaluating risk factors or responses to interventions, obscuring pathogen-specific responses and leading to inconsistent or contradictory conclusions. By analyzing pathogen and reservoir-specific relationships, this study provides a more nuanced understanding of how oral hygiene may influence colonization dynamics. Taken together, these findings highlight the need for targeted research and larger-scale studies that consider pathogen-specific mechanisms in the development of preventive strategies.

## Limitations

We did not characterize pathogen strains, and it is certainly possible that there are strain-specific differences in the likelihood of colonizing different sites. As such, certain strains might be impacted by oral health while the species as a whole may not be. Additional limitations of this study include a limited number of survey participants, high correlation among survey responses reflecting overlapping behaviors, and imbalanced group sizes concerning vaccination status. Finally, our oral health data were collected cross-sectionally, limiting the interpretation to associations rather than causal relationships. Future studies should therefore study oral health behaviors and pathogen carriage longitudinally for greater confidence in these relationships or implement pathogen-specific interventions to establish causality.

## Supporting information

Anonymized data

## Data Availability

Anonymized data that support the findings of this study are included as supporting information.

## Declarations

### Ethics approval and consent to participate

Approval for this project was granted by the Northern Arizona University Institutional Review Board (# 1766728). Participant recruitment for pathogen surveillance began Oct 27th of 2021 and continued until the last sampling event on Oct 31st of 2023. All participants provided formal written consent. Oral health surveys were provided to participants following the last sampling event.

### Consent for publication

Not applicable

### Availability of data and materials

Anonymized data that support the findings of this study are included as supporting information.

### Authors’ contributions

This study was conceptualized by ST, TP, and VYF. Data collection was carried out by CB, AR, TP, and TNF, and formal analyses were performed by RNW and TNF. Writing was completed by RNW, TNF, TP, VYF, and ST. Wet lab work and sample processing were performed by RNW, AR, ST, SM, SW, KD, and KR. All authors have reviewed the manuscript.

## Acknowledgements

We would like to thank the staff at the long-term care facilities for facilitating recruitment and sample collection.

## References

1. Tong S, Amand C, Kieffer A, Kyaw MH. Trends in healthcare utilization and costs associated with pneumonia in the United States during 2008–2014. BMC Health Services Research. 2018;18: 1–8.

2. Loeb M, Carusone SC, Goeree R, Walter SD, Brazil K, Krueger P, et al. Effect of a clinical pathway to reduce hospitalizations in nursing home residents with pneumonia: a randomized controlled trial. JAMA. 2006;295: 2503–2510.

3. Stamm DR, Katta S, Stankewicz HA. Nursing Home–Acquired Pneumonia. StatPearls [Internet]. StatPearls Publishing; 2023.

4. Kang Y-S, Ryoo SR, Byun SJ, Jeong Y-J, Oh JY, Yoon Y-S. Antimicrobial Resistance and Clinical Outcomes in Nursing Home-Acquired Pneumonia, Compared to Community-Acquired Pneumonia. Yonsei Medical Journal. 2016;58: 180.

5. Ewig S, Klapdor B, Pletz MW, Rohde G, Schütte H, Schaberg T, et al. Nursing-home-acquired pneumonia in Germany: an 8-year prospective multicentre study. Thorax. 2012;67: 132–138.

6. Marrie TJ. Pneumonia in the long-term-care facility. Infection control and hospital epidemiology. 2002;23. doi:10.1086/502030

7. Henig O, Kaye KS. Bacterial Pneumonia in Older Adults. Infectious Disease Clinics of North America. 2017;31: 689.

8. Polverino E, Dambrava P, Cillóniz C, Balasso V, Marcos MA, Esquinas C, et al. Nursing home-acquired pneumonia: a 10 year single-centre experience. Thorax. 2010;65: 354–359.

9. Maruyama T, Niederman MS, Kobayashi T, Kobayashi H, Takagi T, D’Alessandro-Gabazza CN, et al. A prospective comparison of nursing home-acquired pneumonia with hospital-acquired pneumonia in non-intubated elderly. Respir Med. 2008;102: 1287–1295.

10. Polverino E, Dambrava P, Cillóniz C, Balasso V, Marcos MA, Esquinas C, et al. Nursing home-acquired pneumonia: a 10 year single-centre experience. Thorax. 2010;65: 354–359.

11. Fernández-Sabé N, Carratalà J, Rosón B, Dorca J, Verdaguer R, Manresa F, et al. Community-acquired pneumonia in very elderly patients: causative organisms, clinical characteristics, and outcomes. Medicine. 2003;82: 159–169.

12. Zalacain R, Torres A, Celis R, Blanquer J, Aspa J, Esteban L, et al. Community-acquired pneumonia in the elderly: Spanish multicentre study. Eur Respir J. 2003;21: 294–302.

13. El-Solh AA, Sikka P, Ramadan F, Davies J. Etiology of severe pneumonia in the very elderly. Am J Respir Crit Care Med. 2001;163: 645–651.

14. Riquelme R, Torres A, El-Ebiary M, de la Bellacasa JP, Estruch R, Mensa J, et al. Community-acquired pneumonia in the elderly: A multivariate analysis of risk and prognostic factors. Am J Respir Crit Care Med. 1996;154: 1450–1455.

15. Vila-Corcoles A, Ochoa-Gondar O, Rodriguez-Blanco T, Raga-Luria X, Gomez-Bertomeu F, EPIVAC Study Group. Epidemiology of community-acquired pneumonia in older adults: a population-based study. Respir Med. 2009;103: 309–316.

16. Rello J, Leeper KV. Severe Community Acquired Pneumonia. Springer Science & Business Media; 2013.

17. Overview of Pneumonia. Goldman’s Cecil Medicine. W.B. Saunders; 2012. pp. 587–596.

18. Kumar V, Abbas AK, Aster JC. Robbins Basic Pathology. Elsevier Health Sciences; 2013.

19. Blain A, MacNeil J, Wang X, Bennett N, Farley MM, Harrison LH, et al. Invasive Haemophilus influenzae Disease in Adults ≥65 Years, United States, 2011. Open Forum Infectious Diseases. 2014;1. doi:10.1093/ofid/ofu044

20. Scannapieco FA, Stewart EM, Mylotte JM. Colonization of dental plaque by respiratory pathogens in medical intensive care patients. Crit Care Med. 1992;20: 740–745.

21. Uno I, Kubo T. Risk Factors for Aspiration Pneumonia among Elderly Patients in a Community-Based Integrated Care Unit: A Retrospective Cohort Study. Geriatrics. 2021;6: 113.

22. Aspiration pneumonia: A renewed perspective and practical approach. Respiratory Medicine. 2021;185: 106485.

23. About Pneumococcal Vaccine: For Providers. 25 Jun 2024 [cited 18 Aug 2024]. Available: https://www.cdc.gov/vaccines/vpd/pneumo/hcp/about-vaccine.html

24. Ben-Shimol S, Regev-Yochay G, Givon-Lavi N, van der Beek BA, Brosh-Nissimov T, Peretz A, et al. Dynamics of Invasive Pneumococcal Disease in Israel in Children and Adults in the 13-Valent Pneumococcal Conjugate Vaccine (PCV13) Era: A Nationwide Prospective Surveillance. Clin Infect Dis. 2022;74: 1639–1649.

25. Ten year public health impact of 13-valent pneumococcal conjugate vaccination in infants: A modelling analysis. Vaccine. 2020;38: 7138–7145.

26. Streptococcus pneumoniae carriage, serotypes, genotypes, and antimicrobial resistance trends among children in Portugal, after introduction of PCV13 in National Immunization Program: A cross-sectional study. Vaccine. 2024;42: 126219.

27. Cleary DW, Jones J, Gladstone RA, Osman KL, Devine VT, Jefferies JM, et al. Changes in serotype prevalence of Streptococcus pneumoniae in Southampton, UK between 2006 and 2018. Scientific Reports. 2022;12: 1–11.

28. Løchen A, Croucher NJ, Anderson RM. Divergent serotype replacement trends and increasing diversity in pneumococcal disease in high income settings reduce the benefit of expanding vaccine valency. Scientific Reports. 2020;10: 1–17.

29. Jung YH, Choe YJ, Lee CY, Jung SO, Lee DH, Yoo JI. Impact of national pneumococcal vaccination program on invasive pneumococcal diseases in South Korea. Scientific Reports. 2022;12: 1–8.

30. Berg R, Berkey DB, Tang JM, Baine C, Altman DS. Oral health status of older adults in Arizona: results from the Arizona Elder Study. Spec Care Dentist. 2000;20: 226–233.

31. Didilescu AC, Skaug N, Marica C, Didilescu C. Respiratory pathogens in dental plaque of hospitalized patients with chronic lung diseases. Clin Oral Investig. 2005;9: 141–147.

32. El-Solh AA, Pietrantoni C, Bhat A, Okada M, Zambon J, Aquilina A, et al. Colonization of dental plaques: a reservoir of respiratory pathogens for hospital-acquired pneumonia in institutionalized elders. Chest. 2004;126: 1575–1582.

33. Person JE. Russell Kirk: A Critical Biography of a Conservative Mind. Madison Books; 1999.

34. Russell SL, Boylan RJ, Kaslick RS, Scannapieco FA, Katz RV. Respiratory pathogen colonization of the dental plaque of institutionalized elders. Spec Care Dentist. 1999;19: 128–134.

35. Bassim CW, Gibson G, Ward T, Paphides BM, DeNucci DJ. Modification of the Risk of Mortality from Pneumonia with Oral Hygiene Care. Journal of the American Geriatrics Society. 2008;56: 1601–1607.

36. Yoneyama T, Yoshida M, Ohrui T, Mukaiyama H, Okamoto H, Hoshiba K, et al. Oral Care Reduces Pneumonia in Older Patients in Nursing Homes. Journal of the American Geriatrics Society. 2002;50: 430–433.

37. Sjögren P, Nilsson E, Forsell M, Johansson O, Hoogstraate J. A systematic review of the preventive effect of oral hygiene on pneumonia and respiratory tract infection in elderly people in hospitals and nursing homes: effect estimates and methodological quality of randomized controlled trials. J Am Geriatr Soc. 2008;56: 2124–2130.

38. Chiang T-C, Huang M-S, Lu P-L, Huang S-T, Lin Y-C. The effect of oral care intervention on pneumonia hospitalization, Staphylococcus aureus distribution, and salivary bacterial concentration in Taiwan nursing home residents: a pilot study. BMC Infectious Diseases. 2020;20: 374.

39. Adachi M, Ishihara K, Abe S, Okuda K, Ishikawa T. Effect of professional oral health care on the elderly living in nursing homes. Oral surgery, oral medicine, oral pathology, oral radiology, and endodontics. 2002;94. doi:10.1067/moe.2002.123493

40. Mylotte JM. Will Maintenance of Oral Hygiene in Nursing Home Residents Prevent Pneumonia? Journal of the American Geriatrics Society. 2018;66: 590–594.

41. Lavigne SE, Forrest JL. An umbrella review of systematic reviews of the evidence of a causal relationship between periodontal microbes and respiratory diseases: Position paper from the Canadian Dental Hygienists Association. Canadian Journal of Dental Hygiene. 2020;54: 144.

42. Marusiak MJ, Paulden M, Ohinmaa A. Professional oral health care prevents mouth-lung infection in long-term care homes: a systematic review. Canadian Journal of Dental Hygiene. 2023;57: 180.

43. Whealy RN, Roberts A, Furstenau TN, Timm S, Maltinsky S, Wells S, et al. Longitudinal prevalence and co-carriage of pathogens associated with nursing home acquired pneumonia in three long-term care facilities. bioRxiv. 2024. p. 2024.12.19.629505. doi:10.1101/2024.12.19.629505

44. Cao Y, Liu C, Lin J, Ng L, Needleman I, Walsh T, et al. Oral care measures for preventing nursing home-acquired pneumonia. The Cochrane Database of Systematic Reviews. 2022;2022: CD012416.

45. [No title]. [cited 17 Apr 2025]. Available: https://www.cdc.gov/nchs/data/npals/2020-RCC-DB454-state-tables-508.pdf

46. Simell B, Auranen K, Käyhty H, Goldblatt D, Dagan R, O’Brien KL, et al. The fundamental link between pneumococcal carriage and disease. Expert Rev Vaccines. 2012;11: 841–855.

47. Camilli R, Vescio MF, Giufrè M, Daprai L, Garlaschi ML, Cerquetti M, et al. Carriage of Haemophilus influenzae is associated with pneumococcal vaccination in Italian children. Vaccine. 2015;33: 4559–4564.

48. Cope EK, Goldstein-Daruech N, Kofonow JM, Christensen L, McDermott B, Monroy F, et al. Regulation of virulence gene expression resulting from Streptococcus pneumoniae and nontypeable Haemophilus influenzae interactions in chronic disease. PLoS One. 2011;6: e28523.

49. Bogaert D, De Groot R, Hermans PWM. Streptococcus pneumoniae colonisation: the key to pneumococcal disease. Lancet Infect Dis. 2004;4: 144–154.

50. Biesbroek G, Wang X, Keijser BJF, Eijkemans RMJ, Trzciński K, Rots NY, et al. Seven-valent pneumococcal conjugate vaccine and nasopharyngeal microbiota in healthy children. Emerg Infect Dis. 2014;20: 201–210.

51. Kielbik K, Pietras A, Jablonska J, Bakiera A, Borek A, Niedzielska G, et al. Impact of pneumococcal vaccination on nasopharyngeal carriage of Streptococcus pneumoniae and Microbiota profiles in preschool children in south East Poland. Vaccines (Basel). 2022;10: 791.

52. Zivich PN, Grabenstein JD, Becker-Dreps SI, Weber DJ. Streptococcus pneumoniae outbreaks and implications for transmission and control: a systematic review. Pneumonia (Nathan). 2018;10: 11.

